# The Economic Burden of KCNT1-Related Disorders in the United States: Insights from Caregiver-Reported and EMR-Derived Data

**DOI:** 10.64898/2026.03.16.26348524

**Authors:** Amanda Abuhl, Brad Bryan, Megan Wright, Allison Rosenberg, Justin West, Sarah Drislane

## Abstract

**Background:** KCNT1-related disorders are a rare, severe neurogenetic disorder associated with early-onset, treatment-resistant seizures and significant developmental comorbidities. Currently there are no treatment-modifying therapeutics for this condition, and the condition necessitates complex, lifelong care that places a profound financial strain on affected families and healthcare systems. However, data quantifying this economic burden is sparse.

**Objective:** To evaluate the annual cost burden of KCNT1-related disorders in the United States using both caregiver-reported expenditures and electronic medical record (EMR) data, providing a comprehensive analysis of direct, indirect, and out-of-pocket expenses.

**Methods:** A retrospective cohort analysis was conducted using two complementary data sources. In 2025, 34 U.S-based. caregivers from the KCNT1 Epilepsy Foundation registry completed a survey capturing insurance status, medical and non-medical expenses, and indirect costs. Separately, EMR data from 49 U.S.-based patients with KCNT1 variants were extracted from the Citizen Health database. Clinical services were mapped to CPT and HCPCS codes, and costs were calculated using Medicare fee schedules and other publicly available datasets.

**Results:** Caregiver-reported data revealed that all respondents possessed some form of insurance coverage, primarily through private insurance purchased independently or through their employer, or Medicaid. Nearly half of respondents (18/34) experienced financial hardship, citing high out-of-pocket expenses, medical debt, and loss of income due to caregiving responsibilities, and twelve percent of respondents delayed treatment due to financial strain (n=4). The estimated mean total annual medical cost per family—including direct, indirect, non-medical, and non-covered expenses—ranged from $355,474 to $797,727, based on upper and lower bounds of response categories from 10 respondents. EMR analysis, which only reported on direct medical costs, revealed that average first-year direct medical costs reached $154,389 per patient based on the records from 49 patients. This cost was primarily driven by hospitalizations, medications, and therapeutic procedures. Based on EMR data, direct medical costs declined once the patients reached two years of age and stabilized in subsequent years. Hospitalizations remained the most substantial cost contributor regardless of the age of the patient.

**Conclusion:** KCNT1-related disorders imposes a substantial economic burden on families and healthcare systems, particularly in the first year after diagnosis. This study highlights the need for rapid diagnostic procedures, targeted therapies, improved insurance coverage, and legislative support for families managing rare, high-burden conditions. Findings provide essential cost data to support drug development, healthcare planning, and rare disease policy reform.

**Significance:** This is the first U.S.-based study to quantify both medical and non-medical costs associated with KCNT1-related disorders using combined caregiver and EMR data. The results highlight the urgency of disease-modifying treatments and equitable access to care, informing clinical trials and advocacy for systemic healthcare support.

## Introduction

KCNT1-related disorders are a rare and severe genetic conditions caused by pathogenic variants in the *KCNT1* gene, which encodes a sodium-activated potassium channel involved in neuronal excitability [1–3]. Pathogenic variants in *KCNT1* result in hyperactive neuronal firing, leading to drug-resistant epilepsy [4]. Among the clinical phenotypes associated with *KCNT1* variants are Epilepsy of Infancy with Migrating Focal Seizures (EIMFS), Early Infantile Developmental and Epileptic Encephalopathy (EIDEE), West syndrome, and sleep hypermotor epilepsy (SHE, formerly known as autosomal dominant nocturnal frontal lobe epilepsy, ADNFLE) [5–6]. These conditions are characterized by frequent, often intractable seizures that significantly impair neurological development and quality of life [5–7].

KCNT1-related disorders are diagnosed through genetic testing [8, 9], often prompted by early-onset or childhood seizures that do not respond to conventional anti-seizure medications [4–7]. Seizure types include focal seizures, tonic-clonic seizures, and epileptic spasms, all of which can be frequent and severe [2,6,11,12]. The disorder is associated with a high rate of comorbidities, including developmental delay, cognitive impairment, and motor dysfunction [2–7]. The severity and frequency of seizures contribute to a high burden of care, necessitating frequent hospitalizations and specialized interventions, with most affected individuals requiring lifelong medical support. Behavioral challenges, including hyperactivity and aggression, may also be present, further complicating patient management [2,6]. The progressive nature of the disorder means that symptoms often worsen over time, requiring continuous adjustments in treatment plans and supportive therapies. Effective treatment options remain limited, and most current interventions focus on symptom management rather than disease modification.

Due to the complex and chronic nature of KCNT1-related disorders, affected individuals require multidisciplinary medical care [2]. This includes regular specialized pediatric neurological evaluations, continuous medication management, genetic counseling, physical and occupational therapy, and, in some cases, surgical interventions such as vagal nerve stimulation (VNS) [13,14]. Additionally, emergency medical care for status epilepticus and other seizure-related complications is common. Many children require assistive devices such as wheelchairs, communication aids, and orthotic supports due to motor impairments. The high frequency of medical visits, specialized treatments, and the need for caregiver support place a significant financial and emotional strain on families. The burden on caregivers is substantial, often necessitating reduced work hours or full-time caregiving, leading to lost income and increased financial distress. In some cases, families must relocate to be closer to specialized medical centers, further adding to economic strain.

The United States healthcare system differs substantially from those of many other high-income countries in that it does not provide universal, government-funded health coverage. Instead, healthcare financing is fragmented across a mix of private and public insurance programs, with coverage often linked to employment [15–18]. Most children and working-age adults are insured through employer-sponsored private insurance, while public programs such as Medicaid (for low-income individuals and children) and

Medicare (primarily for adults aged ≥65 years and individuals with disabilities) provide coverage for specific populations. Despite these programs, gaps in coverage remain, and families frequently face substantial out-of-pocket costs, including insurance premiums, deductibles, copayments, and expenses for services not fully covered by insurance. Access to specialized care, advanced therapies, and durable medical equipment often depends on insurance type, state-level policies, and prior authorization requirements. As a result, families affected by rare and complex conditions may experience significant financial burden even when insured, and healthcare utilization patterns can vary widely across regions and socioeconomic groups. Many studies of rare diseases have tallied the significant healthcare costs and economic strain on families [19–24]. In 2023, US healthcare spending averaged $14,570 per person, but for individuals with a rare disease, annual costs were estimated at $62,084 [25]. While this data represents the overall aggregate spending per person across all rare diseases, the full extent of this financial burden remains unreported for KCNT1-related disorders. Due to the rarity of the condition and the variability in healthcare needs, existing data on the financial impact of KCNT1-related disorders is limited. Direct medical expenses include frequent hospitalizations, specialized medical care, and long-term medication use, and families also incur out-of-pocket expenses for assistive devices, therapies, and other uncovered medical treatments. The indirect costs associated with KCNT1-related disorders, such as lost wages for caregivers, employment limitations, and educational support needs, also remain a poorly characterized aspect of the total cost of KCNT1-related disorders. A comprehensive assessment of these costs is crucial to clarify the true economic challenges faced by families and healthcare systems.

A better understanding of the economic burden of KCNT1-related disorders is essential for informing healthcare policies and advocating for financial assistance programs [26–27]. By comparing parent-reported expenses with electronic medical record (EMR) data from the United States, this study aims to provide a more accurate and holistic estimate of the financial strain imposed by the disorder on US-based families. Identifying discrepancies between these data sources will also help pinpoint gaps in coverage and access to necessary services, ensuring that affected families receive adequate support. From a research and drug development perspective, quantifying the financial burden of KCNT1-related disorders is critical for advancing new treatments [28–32]. Cost data plays a key role in FDA Investigational New Drug (IND) applications, as they provide evidence of unmet medical need and justify investment in novel therapies which can improve the health of affected individuals, and have the potential to drastically reduce the total indirect and direct costs of care. This study will contribute valuable economic data that can support future drug development efforts and guide research into cost-effective interventions, ultimately improving the lives of individuals affected by KCNT1-related disorders.

## Methods

### Caregiver-Reported Data Survey Study Design, Survey Development, Data Collection

In March to April 2025, thirty-four United States-based respondents were recruited from the KCNT1 Epilepsy Foundation’s Contact Registry. Inclusion criteria required that the respondent is a US-based parent and primary caregiver of a child with a KCNT1 genetic diagnosis, belongs to the KCNT1 Epilepsy Foundation patient association, and could provide estimates of costs. The survey asked primary caregivers to estimate annual health insurance coverage and approximate medical costs, non-medical expenses, and indirect costs based on cost brackets including $0, $1-$5,000, $5,001 to $20,000, $20,001 to $50,000, $50,001 to $100,000, $100,001 to $250,000, and more than $250,000 **(Supplemental File 1)**. The cost brackets used in the survey were selected to capture the wide variability in expenses experienced by families affected by KCNT1-related disorders. Because there are no prior studies providing detailed estimates of medical, non-medical, or indirect costs for this rare disease population, we designed the ranges to encompass both lower and extremely high expenditures, reflecting the potential financial burden of a severe, rare, and chronic condition. The brackets increase progressively to allow respondents to indicate approximate costs without requiring precise calculations, while also ensuring that extremely high-cost scenarios—common in rare, medically complex disorders—were adequately represented. This approach balances respondent ease with the need to capture a broad spectrum of financial experiences. In the US, healthcare services and related expenses are typically itemized through bills and explanation of benefits, allowing caregivers to separately track costs for medical appointments, medications, emergency care, and other services. This itemization enabled respondents to provide estimates for Direct, Indirect, Non-Medical, and Miscellaneous Non-Covered Costs. The survey was informed by consultations with the KCNT1 Epilepsy Foundation, caregivers of individuals with KCNT1-related disorders, academic clinical experts, and representatives from pharmaceutical and biotechnology companies with KCNT1 therapies in development, who provided input prior to survey development and reviewed draft survey questions. The survey was conducted by the KCNT1 Epilepsy Foundation using secure Microsoft Forms with privacy compliant data storage. The surveys were conducted anonymously, and no identifiable private information could be linked to the respondents.

Electronic Medical Record (EMR) Data Extraction: Data from 49 US-based patients with KCNT1-variants were obtained from the Citizen Health database [33]. Clinical data included diagnostic codes, hospitalizations, assistive device usage, and prescribed medications. Although the EMRs contained records for patients of all ages, we limited our analysis of EMRs to children aged 6 years or younger due to an obvious incompleteness of records in older patients, which may result from early mortality, changes in healthcare providers, or loss to follow-up.

### Data Extraction

Diagnostic codes were derived by mapping Systematized Nomenclature of Medicine—Clinical Terms (SNOMED CT) entries to corresponding Healthcare Common Procedure Coding System (HCPCS) and Current Procedural Terminology (CPT) codes. For reference, SNOMED codes provide standardized clinical terminology to describe diagnoses and medical concepts, while CPT and HCPCS codes are used in the United States to classify and bill for medical procedures, services, and supplies. Direct costs associated with diagnostic procedures, therapeutic interventions, and hospital admissions were estimated using the Centers for Medicare & Medicaid Services (CMS) Physician Fee Schedule [34]. Pricing information for durable medical equipment (DME), including ankle-foot orthoses (AFOs), wheelchairs, and other assistive devices, was obtained from the Durable Medical Equipment Coding System (DMECS) via the Palmetto GBA website [35]. Current outpatient medication prices were retrieved from publicly accessible data available on GoodRx [36]. Number of days per hospital admittance were obtained, and hospital per diem charges were estimated using 2022 data from the Kaiser Family Foundation [37]. Hospital pricing varied significantly based on the region. For consistent calculations that reflected the national average, pricing in the state of Nebraska was used to determine hospital expenditure costs, as KFF indicated that Nebraska represents the median per capita hospital expenditure in the United States. The Nebraska median daily cost for hospitalization at the time of analysis ($2,879) was used in this report. As the Citizen data did not account for variations in patient acuity, such as differences between intensive care unit (ICU) and general medical floor stays, the reported financial figures more than certainly underestimate the true cost burden for hospitalizations.

### Data Visualization

Microsoft Excel was used to document and organize cost data associated with the management of KCNT1-related disorders. Descriptive statistics and data visualization were performed using Microsoft Excel and GraphPad Prism v10.

## RESULTS

### Caregiver Reported Financial Burden of KCNT1-related disorders

Based on anonymous caregiver-reported surveys, all respondents (n=34) reported having health coverage. This coverage was through private insurance plans purchased independently or through their employer (79%, n=27), Medicaid (41%, n=14), Medicare (9%, n=3), or Children’s Health Insurance Program (CHIP) (3%, n=1) (**Figure 1A)**. Most families utilized a combination of these plans. Over half of respondents (53%, n=18) reported that medical expenses associated with KCNT1-related disorders had caused financial hardship for their family **(Figure 1B)**, while 16 respondents indicated that they had not experienced financial hardship, suggesting that while cost burdens are substantial, their impact varies based on individual circumstances such as insurance type, income level, and access to supportive resources. When asked in an open-ended format about financial challenges, parents reported facing high out-of-pocket costs for medications, medical supplies, and specialized equipment, and often having to choose between paying medical bills or essential living expenses like groceries and rent. They also described ongoing medical debt, gaps in insurance, lost income from caregiving, and spending significant time and energy fighting for coverage, making it difficult to manage finances and plan for long-term needs. When asked whether cost concerns had ever led their family to delay or avoid seeking medical care for their dependent with a KCNT1-related disorder, 12% (n=4) reported that they had indeed delayed or avoided medical care due to the financial burden of the condition **(Figure 1C)**.

**Figure 1.**
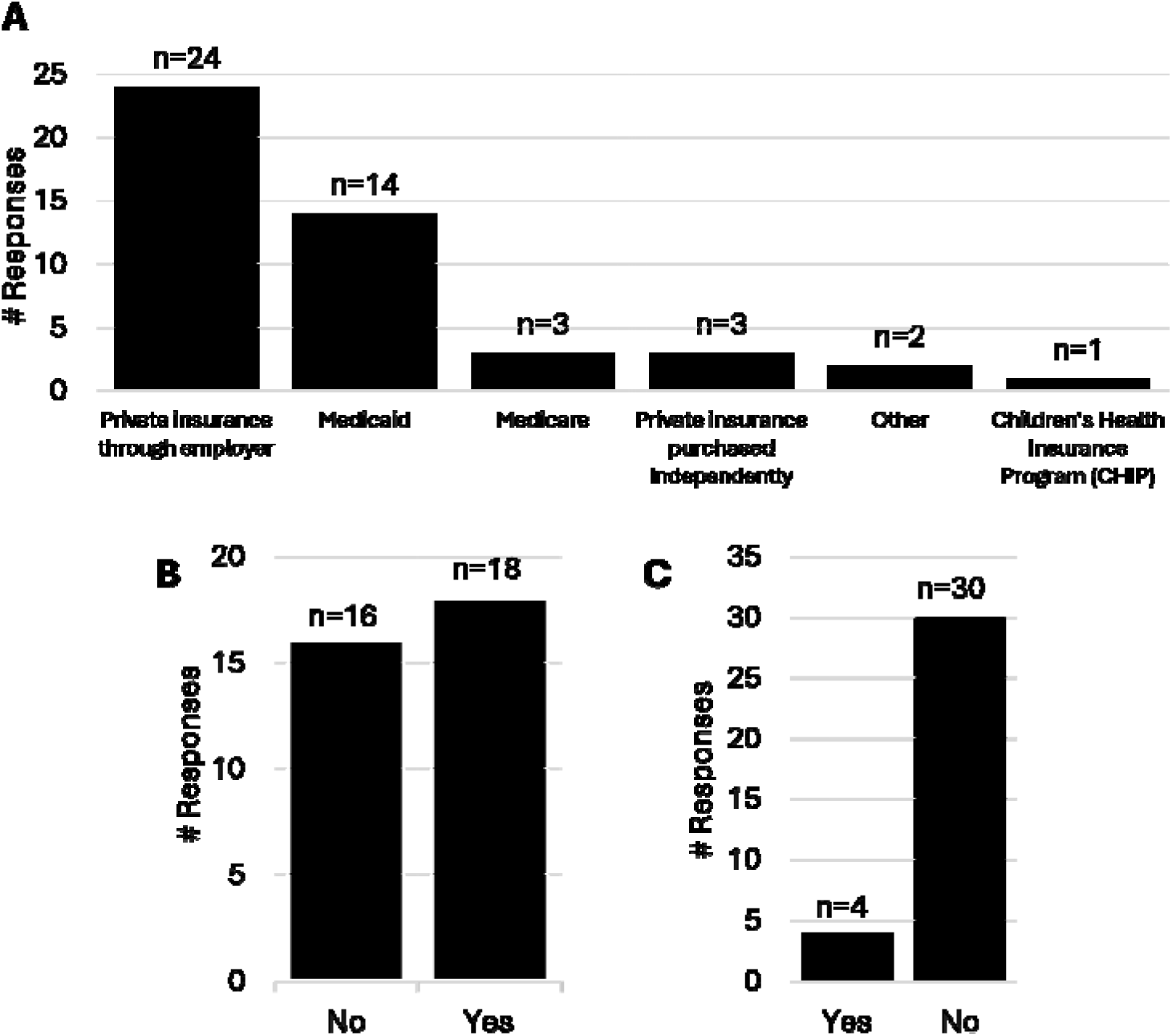
Caregiver-reported financial burden of KCNT1-related disorders. Data collected from a survey of 34 United States-based respondents who served as the primary caregiver for an individual with a KCNT1 variant. All responses are self-reported and completely anonymous. (A) Health insurance coverage status of respondents. Caregivers were given the option to elect all applicable forms of insurance they utilized. (B) Caregiver responses on experiencing financial hardship due to medical expenses. (C) Caregiver responses on delaying or avoiding medical care due to cost concerns.

Of the 34 total respondents, 11 of these provided the annual cost estimate of KCNT1-related financial expenditures across a wide range of medical cost categories (Direct, Indirect, Non-Medical, and Miscellaneous Non-Covered Costs) related to the management of KCNT1-related disorders **(Figure 2)**. Annual direct costs, including core components of ongoing disease management (medical appointments, hospitalizations, medications, medical procedures, and emergency care) were reportedly the most expensive components of annual medical care for patients with a KCNT1-related disorder **(Figure 2A)**. Additionally, families frequently incurred high expenses for home healthcare services, medical supplies, diagnostic tests, and therapies such as physical or occupational rehabilitation. These findings highlight the multifaceted nature of direct medical costs, reflecting the complexity of the disorder and the broad spectrum of care required to address both primary symptoms and secondary health complications. In assessing annual indirect medical costs related to KCNT1-related disorders, informal caregiving emerged as the largest reported expense **(Figure 2B)**. Informal caregiving entails providing daily, unpaid support, which may significantly impact the respondents’ ability to maintain full-time employment or pursue career advancement. In addition to caregiving responsibilities, both job loss and absenteeism due to medical appointments, hospitalizations, or recovery periods contributed meaningfully to the financial burden. These findings highlight the far-reaching economic impact of KCNT1 disorders, extending beyond healthcare expenses to include substantial productivity losses and long-term employment disruptions for affected families. Annual non-medical costs related to KCNT1-related disorders represented a substantial financial burden for many families, with the most significant expenditures reported in nutritional support, special equipment, professional caregiving services, and home modifications to create adaptive living environments **(Figure 2C)**. These expenses reflect the need for ongoing lifestyle adjustments and specialized support that go beyond direct healthcare services. While essential, categories such as educational expenses and psychosocial support—though still reported—accounted for the lowest expenditures in this group. The annual healthcare costs not covered by insurance related to KCNT1-related disorders were generally low **(Figure 2D)**. Over the counter medications accounted for the largest portion of these out-of-pocket costs, followed by alternative/complementary therapies.

**Figure 2.**
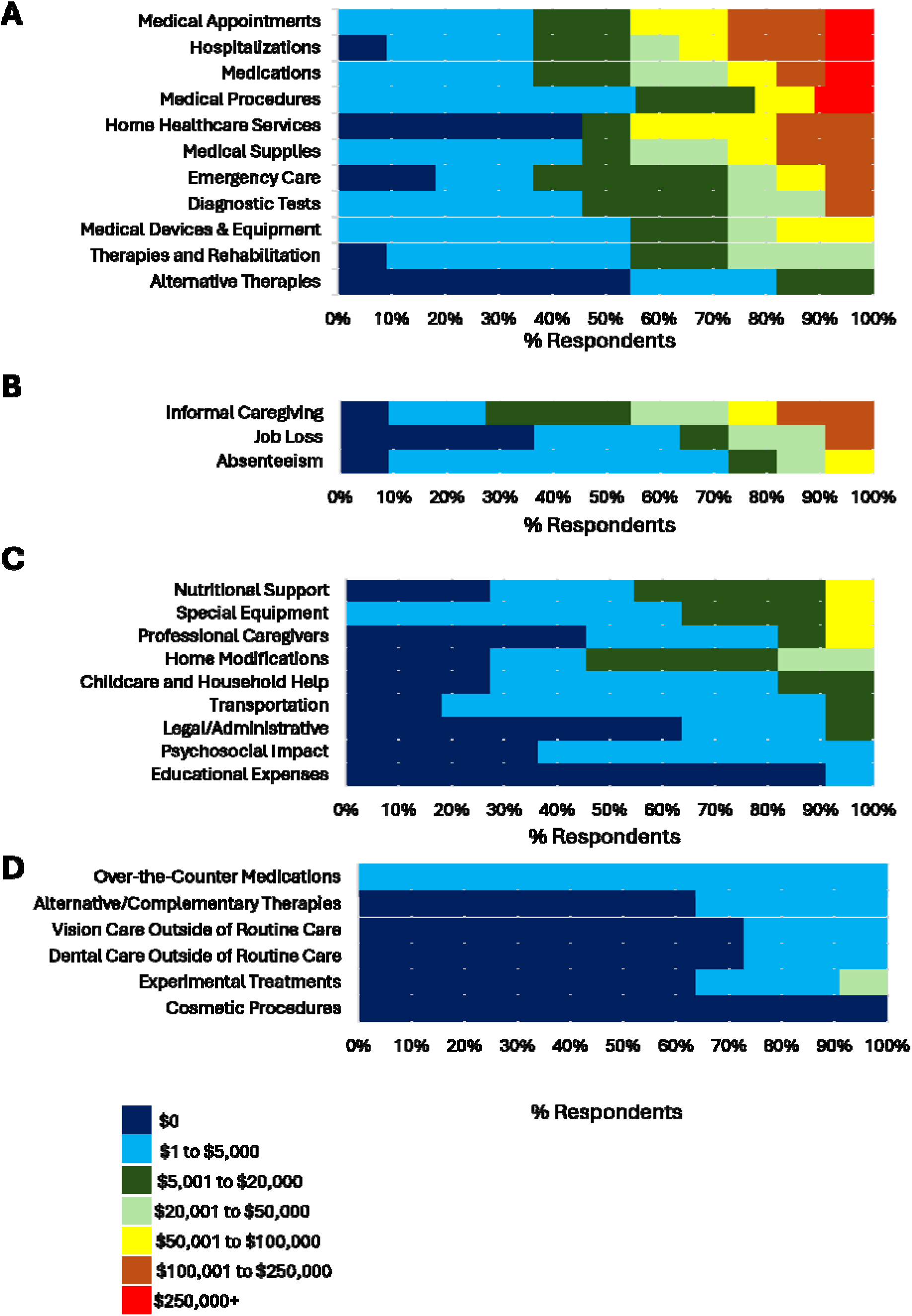
Caregiver-reported financial burden of KCNT1-related disorders was collected from 11 United States-based respondents who served as the primary caregiver for an individual with a KCNT1 variant. To gather information on financial impact, the survey asked caregivers to estimate their expenses by choosing from a set of cost ranges rather than reporting exact amounts. Respondents were asked to provide their annual expenses or loss of income in annual **(A)** Direct Medical Costs (related to the medical management of KCNT1-Related Disorder and its associated symptoms, typically including expenses directly related to diagnosis, treatment, and management, including costs for secondary diagnoses or symptoms stemming from the condition), **(B)** indirect medical costs (productivity loss due to caregiving responsibilities for the affected individual, and the unpaid labor of caregiving), **(C)** non-medical costs (arise from the need to adapt living arrangements, manage daily life challenges, and provide additional support beyond medical care), and **(D)** healthcare costs not generally covered by insurance (widely depending on the specific insurance plan and the nature of the rare disease).

We calculated the total annual care costs for the 11 individuals completing the self-reported survey, demonstrating a profound cost to each of the families **(Figure 3)**. Because respondents reported their costs using predefined brackets rather than exact figures, the totals could only be estimated within a range. To calculate overall financial impact, each selected bracket was assigned both a lower and upper bound, allowing researchers to sum costs across all categories using these limits. This method produced a minimum and maximum estimate of total expenses, rather than a single definitive amount. While this approach offers a practical way to quantify costs from bracketed data, it inherently introduces a degree of uncertainty, as the actual values reported by individuals may fall anywhere within the selected ranges. Estimated minimum annual costs ranged from approximately $5,013 to $1,250,019, while maximum projected costs spanned from $80,000 to $2,375,000. Most responses fell below $150,000 on the low end and under $1.4 million on the high end. The mean annual cost ranged from $355,474 (minimum) to $797,727 (maximum), with the median falling between $265,018 and $620,000. This high cost highlights the substantial financial burden associated with care of an individual with a KCNT1 variant, particularly when compared to the average health spending in the United States for 2023, according to the Centers for Medicare and Medicaid Services, was $14,570 per person [38].

**Figure 3.**
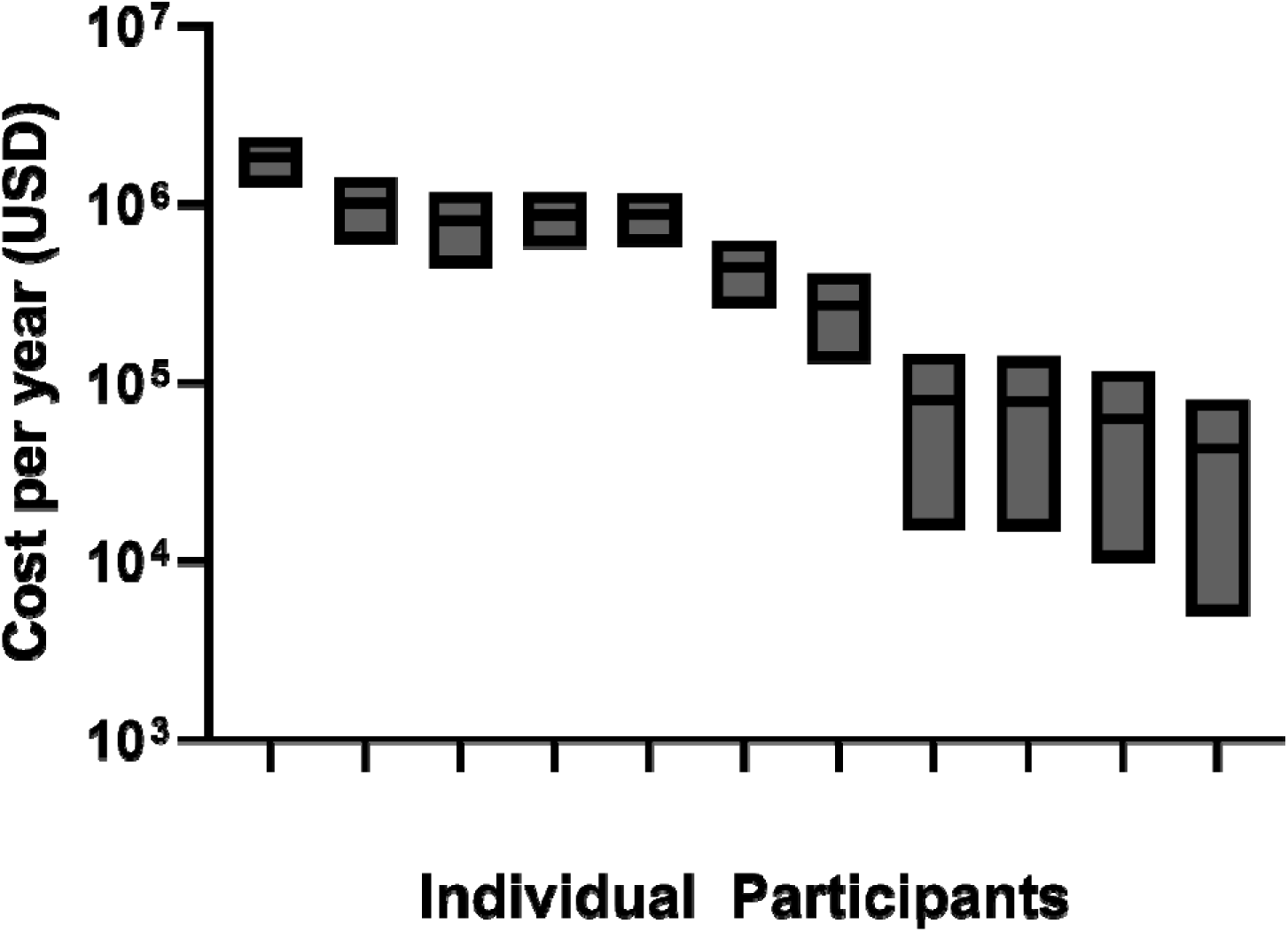
Total annual care costs for individuals with KCNT1-related disorders. The use of cost brackets allowed researchers to calculate an estimated range for the total reported expenses across all caregivers. Since respondents selected from predefined ranges rather than providing exact dollar amounts, the data could only be summarized as minimum and maximum possible totals. For each caregiver, the lowest value of a selected bracket was used to calculate a conservative estimate, while the highest value (or a defined upper limit for open-ended brackets) was used to calculate the maximum estimate. As a result, the aggregated cost data represent a range rather than a precise figure, reflecting the inherent uncertainty introduced by the bracketed response format. This figure presents the calculated total annual care costs for each of the 11 respondents based on self-reported survey data. Given that respondents selected cost brackets for each survey answer, these bars represent the minimum and maximum annual costs per patient.

### Electronic medical record-based analysis of KCNT1-related disorder financial burden

We conducted a retrospective analysis of EMR data using the Citizen Health database to characterize the longitudinal healthcare utilization and associated costs in patients diagnosed with KCNT1-related disorders. Utilizing de-identified EMR data, we extracted information on hospitalizations, medications, durable medical equipment and devices, as well as therapeutic and diagnostic procedures. Our objective was to quantify the direct medical costs incurred over time and to identify patterns in healthcare resource utilization following a KCNT1 diagnosis. This approach aims to inform clinical management strategies and support the development of targeted interventions for this patient population.

The frequency of diagnostic procedures, hospital admissions, and medication prescriptions was highest during the first year of life **(Figures 4A–C)**, marking a period of intense medical intervention. Following this initial peak, all three categories experienced a sharp decline of over 50% in the second year and continued to decrease more gradually in the years that followed. In contrast, the number of therapeutic procedures remained relatively stable across the entire six-year observation period **(Figure 4D)**, suggesting a consistent need for ongoing therapeutic support. Notably, the use of medical devices showed a different pattern, with utilization peaking in the second and third years of life before tapering off in subsequent years **(Figure 4E)**.

**Figure 4.**
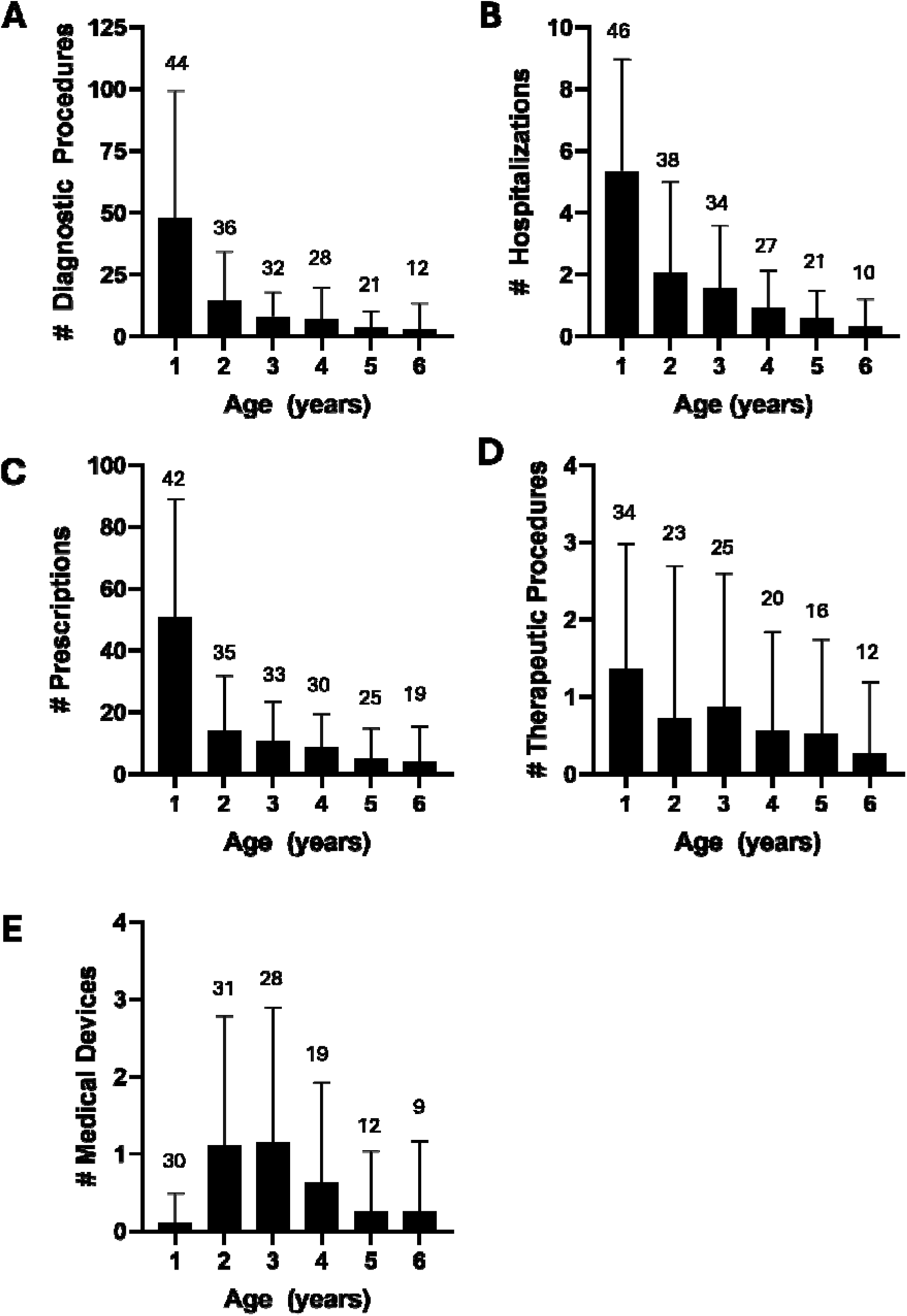
Frequency of healthcare resource utilization based on electronic medical record (EMR) data. This figure illustrates the age-specific frequency of healthcare utilization over the first six years of life, including the average number of **(A)** diagnostic procedures, **(B)** hospital admissions, **(C)** medication prescriptions, **(D)** therapeutic procedures, and **(E)** medical devices used by patients at each year of age. Data are shown for all individuals, with error bars representing the standard deviation. Numbers above the bars indicate the number of KCNT1 patients utilizing each category. Data was derived from the Citizen Health database for patients with KCNT1-variants.

Over the full six-year study period, hospital stays consistently accounted for the largest share of annual medical expenses, underscoring their significant impact on the overall financial burden **(Figure 5A)**. Hospitalization costs declined notably after the first year, serving as the primary factor driving the overall reduction in healthcare expenses over time. Medications and diagnostic procedures were particularly prominent in the first year, reflecting the high intensity of care typically required during infancy. Beginning in year two, the use of medical devices rose substantially, contributing to a shift in the composition of non-hospital-related expenditures. This evolving cost landscape highlights the dynamic nature of pediatric care needs and the critical role hospitalization plays in the financial trajectory of affected families. When analyzing the cumulative healthcare costs incurred during the first year of life, EMR data revealed that the average total expenditure per patient—including hospitalizations, diagnostic procedures, medications, therapeutic procedures, and medical devices—was approximately $154,389 (SD = $117,716; median = $131,997) **(Figure 5B)**.

**Figure 5.**
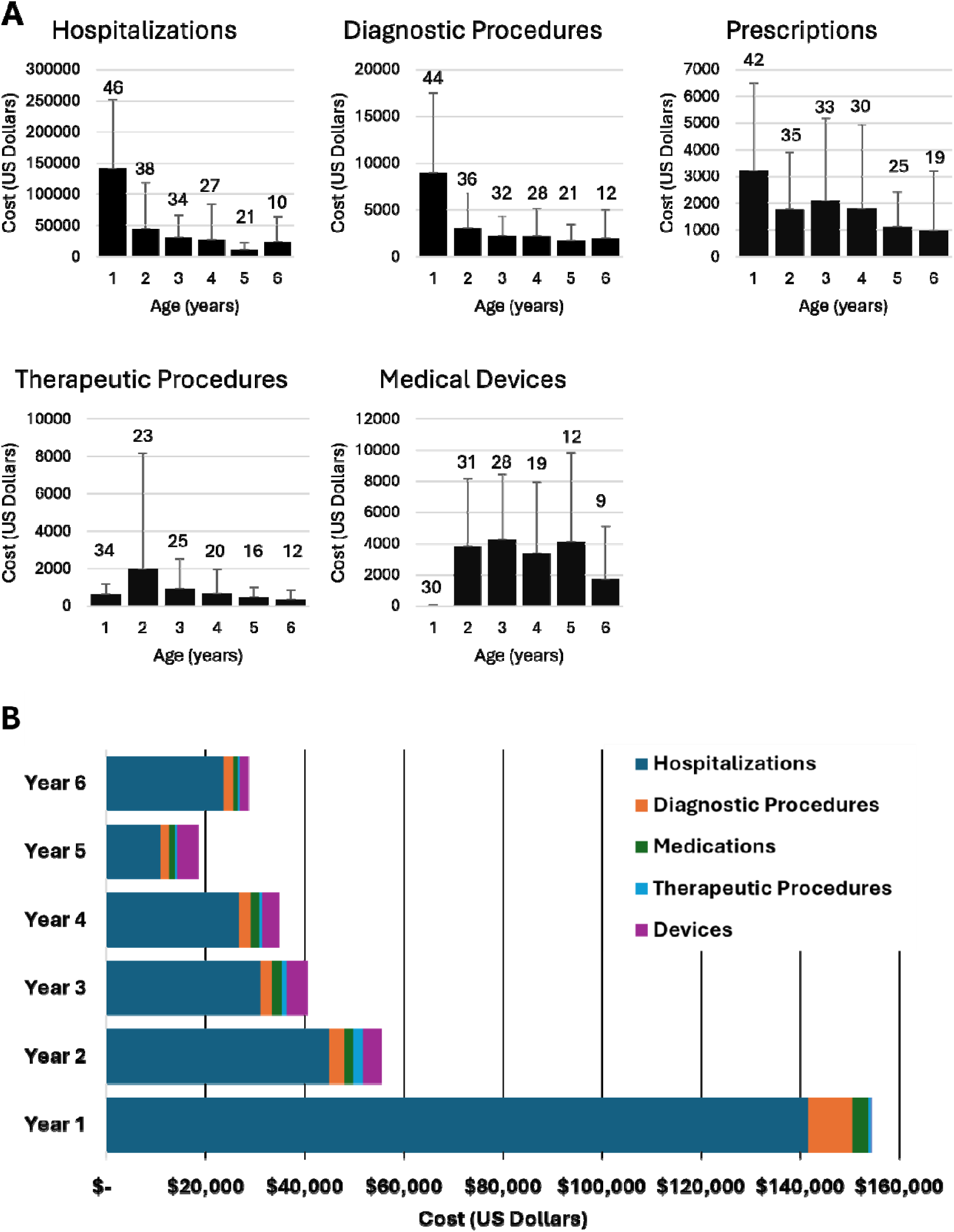
Longitudinal healthcare costs based on electronic medical record (EMR) data from the Citizen Health database for patients with KCNT1-related disorders. (A) The average cost in US Dollars spent for each category of healthcare utilization over the first six years of life. Error bars represent the standard deviation; numbers above the bars indicate the number of KCNT1 patients represented in each year of life. (A) The average annual medical expenses by category were summed for each age of life. Categories of healthcare expenses are divided by color coding. Data was derived from the Citizen Health database for patients with KCNT1-variants.

## DISCUSSION

This study provides a comprehensive assessment of the multifaceted economic burden experienced by families managing KCNT1-related disorders, integrating both caregiver-reported data and electronic medical record (EMR) analyses. Our findings demonstrate the substantial financial impact of this rare neurological disorder, highlighting the necessity for targeted interventions and reforms to alleviate caregiver strain and enhance care accessibility.

The average health spending in the United States for 2023, according to the Centers for Medicare and Medicaid Services, was $14,570 per person, and a national study estimated the average annual cost per patient with a rare disease to be $62,084 [25].

Our data from both caregiver-reported and EMR records show that the financial burden of KCNT1-related disorders far exceeds the average annual cost typically incurred by patients with other rare diseases. Our analysis highlights the limitations of each data source in capturing the full economic burden of KCNT1-related disorders. EMR data indicate that average first-year medical costs are approximately $154,000, with costs declining sharply in subsequent years. In contrast, caregiver-reported annual costs, captured via anonymous surveys using cost brackets, ranged from a mean of $355,474 to $797,727. This higher estimate reflects additional financial burdens not captured in the EMR, including indirect medical costs, non-medical expenses, and other out-of-pocket costs incurred by families. The anonymity of the caregiver reported survey prevents precise age-based comparisons, but the substantial difference between the datasets illustrates that EMR-derived medical costs alone underestimate the true financial impact of KCNT1 on families. Collectively, these findings emphasize the importance of integrating multiple data sources to comprehensively assess the total economic burden of rare, complex disorders.

The substantial direct medical costs observed, particularly in the first year of a patient’s life, align with trends seen in other rare pediatric disorders [39–45]. Our analysis revealed a similar trend for the high cost of direct medical costs, with hospitalizations constituting the largest proportion of annual costs, followed by diagnostic procedures, medications, durable medical equipment, and therapeutic procedures. Notably, the second year of the KCNT1 patients’ life saw a marked decrease in total expenditures, suggesting a shift from intensive diagnostic procedures and in-patient hospital interventions to long-term supportive care. This shift necessitates ongoing financial and logistical support, emphasizing the continued importance of healthcare coverage that extends beyond initial treatment phases.

Caregiver-reported data further illuminates the broader economic impact not necessarily accounted for in EMR data by encompassing indirect and non-medical costs. 18 out of 34 (53%) caregivers reported financial hardship due to out-of-pocket medical expenses, lost income, and other non-medical expenditures. These findings are consistent with broader surveys indicating that a significant proportion of rare disease caregivers experience financial strain, with many reporting the need to reduce work hours or discontinue employment altogether [40–45]

The economic burden of KCNT1-related disorders is consistent with that of other rare pediatric neurological disorders [46–48]. For example, studies on Dravet syndrome and Rett syndrome have documented similar patterns of high initial medical costs followed by a stabilization phase, with caregivers reporting significant indirect costs and financial hardship [49]. These parallels suggest that the economic challenges faced by families managing rare neurological disorders are widespread and not unique to KCNT1-related disorders.

Despite the insights provided, our study has several limitations that must be acknowledged. We acknowledge that the small number of caregivers reporting financial expenditures (11 of 34 total) may limit the generalizability of these results. This was possibly due to privacy concerns or lack of detailed records, which is common in studies of ultra-rare diseases where sample sizes are inherently small. The EMR data utilized did not capture certain costs, including indirect medical costs such as informal caregiving, job loss, or absenteeism, non-medical costs such as nutritional support special equipment, and others, as well as non-covered costs such as over-the-counter medication and other optional treatments. This is strongly reflected in the significantly higher costs reported in the caregiver-reported surveys compared to that pulled from EMR records. Additionally, the caregiver-reported survey may have been subject to recall bias, potentially limiting the generalizability of the findings. Furthermore, both the caregiver reported surveys and the EMR dataset lacked detailed information on the severity of the disorder, which could influence healthcare utilization and associated costs. While we employed average national costs as a reference for cost estimation, regional variations in healthcare expenses may affect the accuracy of our financial assessments. For example, the EMR-based cost model relies on Nebraska hospital per diem rates to approximate hospitalization costs. This may underestimate costs in major pediatric epilepsy centers, which are typically in high-cost urban regions.

Importantly, these data are not generalizable outside the United States, given the location of all participants restricted to within the United States. Finally, because the caregiver reported data was fully anonymous, it lacked demographic and genotypic/phenotypic data which might influence healthcare spending.

Looking forward, our findings advocate for a multifaceted approach to support families affected by rare diseases. Future research should focus on longitudinal studies to track the evolving economic burden over time and evaluate the effectiveness of interventions aimed at reducing caregiver strain and improving care outcomes. Additionally, exploring the experiences of caregivers in different cultural and healthcare settings could provide valuable insights into the universal and unique challenges faced by families managing rare diseases. While this study focuses on the United States, it is important to recognize that healthcare financing models vary widely across countries, and these differences can profoundly affect both direct and indirect cost burdens for families affected by rare conditions such as KCNT1-related disorders. International evidence suggests that rare diseases impose substantial economic burdens globally, with estimates showing high per-patient costs and significant societal impacts beyond direct medical expenses [50–52]. Future work should extend our findings by systematically comparing the financial burden of KCNT1 across international health systems, accounting for differences in insurance coverage, out-of-pocket payment structures, and social support programs, to inform global policy and assistance strategies.

In conclusion, this study demonstrates the significant and multifaceted economic burden of KCNT1-related disorders, highlighting the urgent need for comprehensive support systems that address the diverse needs of affected families. By integrating caregiver perspectives with clinical data, we can inform policies and practices that alleviate the financial and emotional strain on families, ultimately improving the quality of life for both patients and their caregivers.

## Data Availability

The electronic medical record data obtained from Citizen Health are available upon reasonable request by contacting Citizen Health directly. All non Citizen Health data used in this study are provided as supplementary materials to this article.

## Declarations

### Ethics approval and consent to participate

KCNT1 Community Surveys were conducted under approval of a study protocol through Genetic Alliance IRB. Participants were consented, and all survey data were collected anonymously, with no identifying information obtained that could be linked to individual participants. No information regarding medical conditions, genotypes, phenotypes, or disease progression was collected. EMR analysis was reviewed by Pearl IRB and determined to be exempt from Institutional Review Board (IRB) oversight under the U.S. Department of Health and Human Services regulations at 45 CFR 46.104(d), Category 2. Electronic health record (EHR) data obtained from Citizen Health were evaluated and determined to meet the criteria for Non-Human Subjects Research (NHSR). This study was conducted in accordance with the principles of the Declaration of Helsinki.

### Consent for publication

Not applicable.

### Availability of data and materials

The electronic medical record data obtained from Citizen Health are available upon reasonable request by contacting Citizen Health directly. All non–Citizen Health data used in this study are provided as supplementary materials to this article.

## Authors’ contributions

AA contributed to the study design, collected and analyzed the data, and wrote the manuscript.

BB contributed to the study design, collected and analyzed the data, and wrote the manuscript.

MW collected the data.

AR contributed to the study design and wrote the manuscript.

JW contributed to the study design.

SD contributed to the study design and collected the data.

## Acknowledgements

We gratefully acknowledge Citizen Health for partnering with the KCNT1 Epilepsy Foundation to collect, annotate, and make available patient medical records.

