## Supplemental File for "The Economic Burden of KCNT1-Related Disorders in the United States: Insights from Caregiver-Reported and EMR-Derived Data"

**Supplemental File 1. Anonymous survey on insurance and healthcare costs for KCNT1-Related Disorders.**

1) Do you currently have health insurance?

Yes

No

2) If yes, what type of health insurance do you have? (Select all that apply)

Private insurance through employer

Private insurance purchased independently

Medicare

Medicaid (due to financial qualifications)

Medicaid (for a child or dependent on a waiver due to medical/developmental disability)

Children’s Health Insurance Program (CHIP)

Other

3) Have medical expenses caused financial hardship for you or your family?

Yes

No

4) Have you ever delayed or avoided seeking medical care due to cost concerns?

Yes

No

5) What are your annual Direct Medical Costs (Covered by Insurance) related to the management of KCNT1-Related Disorder and its associated symptoms? Direct Medical Costs typically include expenses directly related to the diagnosis, treatment, and management of KCNT1-Related Disorder, including costs for secondary diagnoses or symptoms stemming from the condition (e.g., neurology, respiratory issues, gastrointestinal care, etc.).

**Category**

Medical Appointments

Medications

Diagnostic Tests

Medical Procedures

Medical Supplies

Medical Devices and Equipment

Hospitalizations

Emergency Care

Therapies and Rehabilitation

Home Healthcare Services

Alternative Treatments

Answer choices: $0, $1–$5,000, $5,001–$20,000, $20,001–$50,000, $50,001–$100,000, $100,001–$250,000, and over $250,000

6) What are your current annual Indirect Medical Costs pertaining to the KCNT1-Related Disorder Indirect Medical Costs including productivity loss due to caregiving responsibilities or the rare disease itself.

**Category**

Absenteeism: Daily Salary * Days Absent per Year due to medical appointments, treatments, hospitalizations, or recovery periods

Job Loss: Daily Salary * Days Unable to Work per year due to permanent or temporary loss of employment, reduced work hours, or career changes.

Informal Caregiving: Days spent providing unpaid care, which may lead to reduced work hours or career interruptions.

Answer choices: $0, $1–$5,000, $5,001–$20,000, $20,001–$50,000, $50,001–$100,000, $100,001–$250,000, and over $250,000

7) What are your annual Non-Medical Costs pertaining to the KCNT1-Related Disorder? Indirect Non-Medical Costs are a wide range of expenses that are not directly associated with medical treatments or healthcare services. These costs often arise from the need to adapt living arrangements, manage daily life challenges, and provide additional support beyond medical care.

**Category**

Special Equipment

Transportation to medical appointments

Hiring professional caregivers

Educational Expenses (tutoring, special education, home schooling)

Nutritional Support

Home modifications for adaptive living spaces to accommodate the needs of the patient.

Childcare and Household Help

Psychosocial Impact: Cost associated with mental health support, counseling, therapy, etc.

Legal and Administrative Expenses

Answer choices: $0, $1–$5,000, $5,001–$20,000, $20,001–$50,000, $50,001–$100,000, $100,001–$250,000, and over $250,000

8) What are your annual Healthcare Costs Not Covered by Insurance pertaining to the KCNT1-Related Disorder? Healthcare costs not covered by insurance vary widely depending on the specific insurance plan and the nature of the rare disease.

**Category**

Experimental Treatments (Clinical Trials or accessing therapies not approved by health insurance providers or regulatory agencies)

Alternative or Complementary Therapies

Over-the-Counter Medications

Dental Care outside of routine care

Vision Care outside of routine care

Cosmetic procedures

Answer choices: $0, $1–$5,000, $5,001–$20,000, $20,001–$50,000, $50,001–$100,000, $100,001–$250,000, and over $250,000
